# A prospective observational cohort study to investigate the effect of frailty on mortality of adults following lower limb amputation

**DOI:** 10.1101/2023.07.28.23293258

**Authors:** S Welsh, J Gale, B Carter, K Hussey, D Orr, T Quinn, P Braude

## Abstract

**Introduction:** Frailty is common in patients with atherosclerotic disease and is associated with substantially increased morbidity, mortality and significant economic and resource implications. Major limb amputation (MLA) secondary to critical limb threatening ischaemia (CLTI) is also associated with high mortality rates. This study aims to examine the association between frailty, as defined by the Rockwood Clinical Frailty Scale (CFS), on mortality rates in patients undergoing MLA for CLTI.

**Methods:** This multi-centre, prospective observational cohort study will collect data on MLA performed for CLTI between November 2017 to December 2021 (North Bristol Trust) and January 2016 to October 2021 (NHS Greater Glasgow & Clyde). All patients undergoing MLA for CLTI will be included. Exclusion criteria are MLA for other aetiology, insufficient data to generate CFS score and minor lower limb amputations. Data collected includes age, sex, deprivation index and Charlson Comorbidity Index variables. A consultant/registrar with specialist-interest in frailty will allocate CFS scores based on pre-operative functional status. Three categories will be used: robust (CFS 1-3), mildly frail (CFS 4-5) and frail (CFS 6-8). CFS 9, ‘terminally ill’, will be presented separately. Primary outcome is all-cause mortality following MLA. Secondary outcome is length of stay. Sample size calculation assumed a mortality of 30% in robust and 40% in frail patients (HR=0.7) in a 1:2 ratio (robust:frail), calculating 1000 patients required, using a 0.05 significance level and 90% power. Outcome data will be analysed by multivariable Cox proportional baseline hazards regression controlling for demographic and operative variables (e.g., sex, age, deprivation index, comorbidity index, urgency of operation).

**Discussion:** It is expected the study results will inform clinical decision-making and contribute toward an evidence pool which will inform service planning.

**KEY STUDY CONTACTS:** 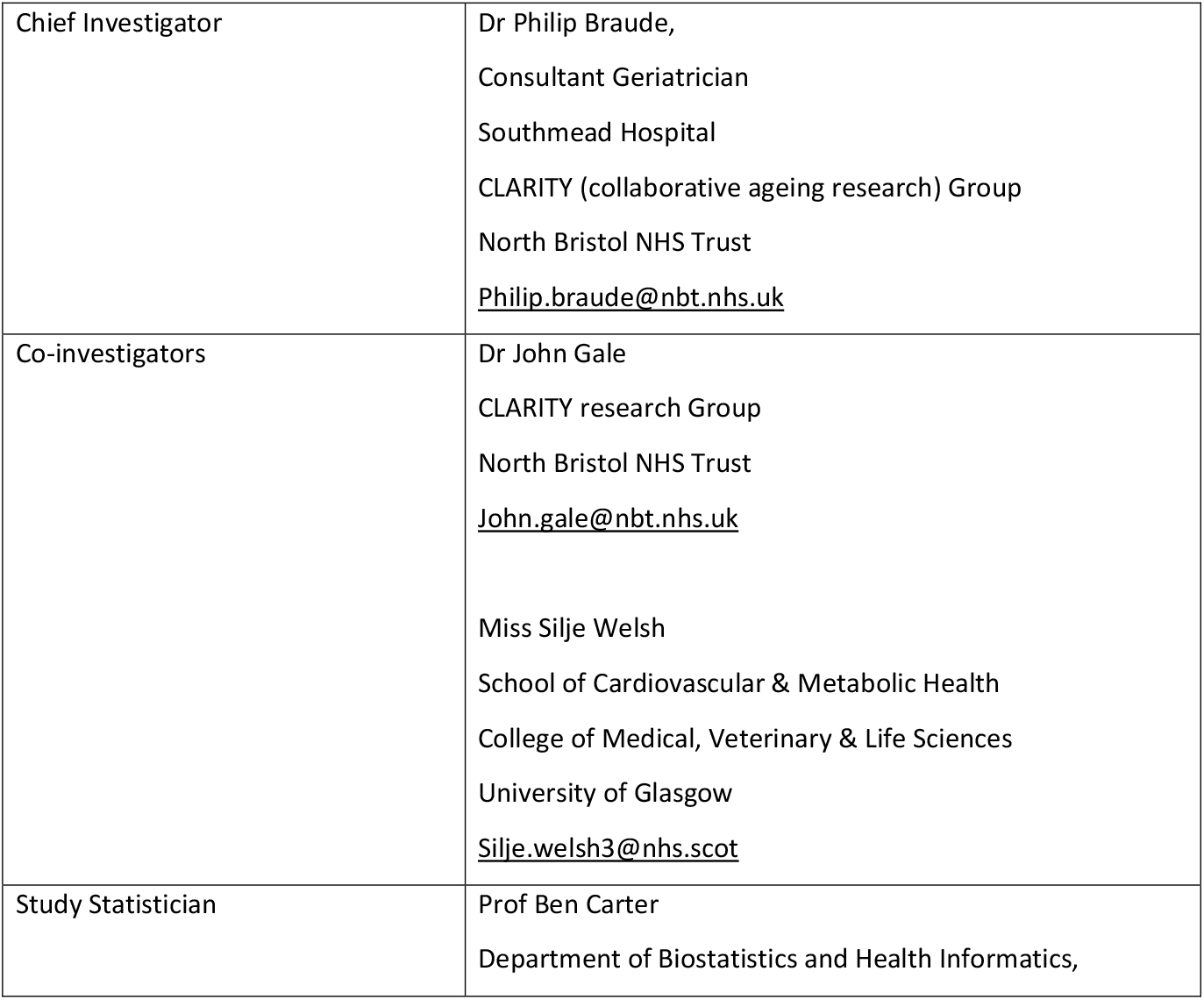

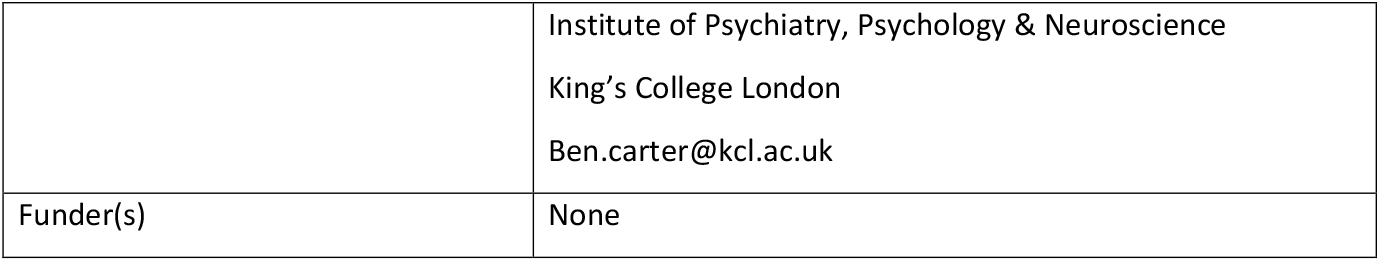

**STUDY SUMMARY:** 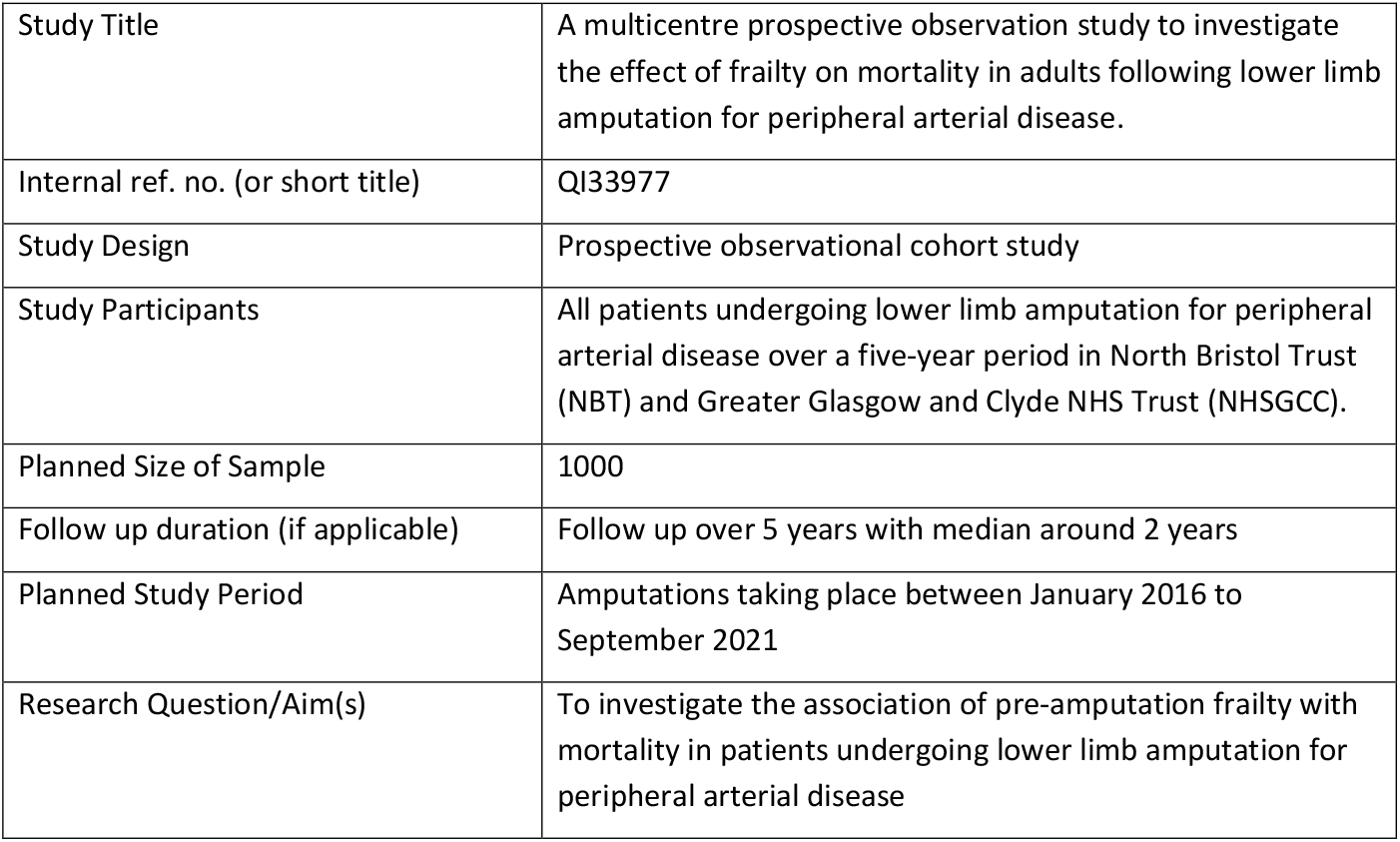

**FUNDING AND SUPPORT IN KIND:** 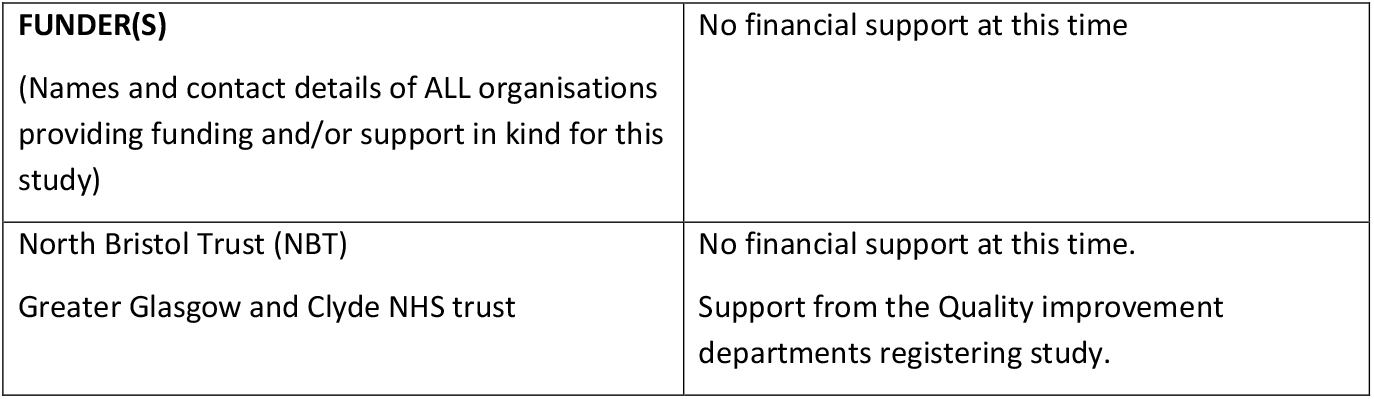

**PROTOCOL CONTRIBUTORS:** 

**Study contributors:**

1. NBT vascular department – proving data on all patients that have undergone LLA in the last 5 years.
2. NHSGGC vascular department – proving data on all patients that have undergone LLA in the last 5 years.

## 1 BACKGROUND

Frailty is a multi-faceted change in a person’s functional status and physiological reserve and is a common clinical syndrome in older adults, carrying an increased risk for poor health outcomes including falls, incident disability, hospitalisation and mortality. It has been shown that frailty can predict recovery and outcomes in trauma patients. It is also an independent predictor of 30-day mortality, delirium and increased care requirement. Previous studies have found that frailty is associated with poorer perioperative outcomes and 30-day mortality in patients undergoing major limb amputation (MLA). This is the first study aiming to investigate the association between MLA and frailty, as defined by the Rockwood Clinical Frailty Scale (CFS)^(1)^, on a large scale.

Chronic limb threatening ischaemia (CLTI) represents end stage atherosclerotic peripheral arterial disease and is the most common cause for MLA of the lower limbs in the UK. CLTI is associated with high mortality with some estimates of 1-year mortality rates upwards of 50%^(2)^. As atherosclerotic disease is systemic, it is often associated with concomitant co-morbidity, with inherent physiological burden, such as ischaemic heart disease, chronic kidney disease and cerebrovascular disease, contributing toward the observed significant mortality rates.

Cardiovascular disease is associated with increased risk of frailty. Clinical frailty scoring based on functional and cognitive status is becoming increasingly validated as a tool for risk stratifying patients. This multicentre study will examine how varying levels of frailty (defined by the Clinical Frailty Scale) are associated with mortality in patients undergoing MLA for CLTI. This will allow for better shared decision-making around MLA, targeted review for high-risk patients and improved service planning.

## 2 RESEARCH QUESTION

### 2.1 Objectives

To investigate whether the frailty (as assessed by the CFS prior to admission to hospital) is associated with mortality in adults undergoing lower limb amputation for peripheral arterial disease.

### 2.2 Outcome

Primary Outcome: All-cause mortality from admission for amputation. Patient who are not contactable due to moving away from the hospital catchment will be censored at the point last known alive.

Secondary outcomes: Length of index hospital stay. Patients will be censored at the point of mortality.

## 3 STUDY DESIGN and METHODS of DATA COLLECTION

We are conducting a prospective observational cohort study. Our data collection process is outlined in the following steps:

We will collect data on all patients who have had a lower limb amputation in North Bristol Trust and the Greater Glasgow and Clyde NHS Health Board over a 5-year period (2016-2021). We will then screen all patients for the indication for amputation, excluding those who have had amputations for other reason than PAD.

A Consultant or Registrar with training in frailty assessment will retrospectively allocate a CFS score based on their pre-operative functional status documented in electronic/paper healthcare records. The use of CFS to retrospective assess frailty is a validated technique^(3)^.

We will use a prospective ‘time to event analysis’. Follow-up is defined as ‘last known alive’ – judged by the patient’s last point of contact with medical services (e.g., outpatient or inpatient medical assessment, blood tests or collection of medical prescriptions).

## 4 STUDY SETTING

NBT is a vascular tertiary referral centre carrying out around 60 – 80 lower limb amputations per year. NHSGGC is also a vascular tertiary referral centre which carries out between 100-120 amputations annually. As this is a multicentre study results will be more generalisable to the UK population.

## 5 SAMPLE AND RECRUITMENT

### 5.1 Eligibility Criteria

The study sample will include all patients admitted from November 2017 until December 2021 at NBT and January 2016 to October 2021 at NHSGCC for a lower limb amputation (elective and/or emergency).

#### 5.1.1 Inclusion criteria

All patients who have had an MLA for CLTI over the study period, including transtibial, transfemoral and through the knee amputations. Indication will be judged by discharge summary, operation note or electronic medical records. Patients with mixed aetiology indication (e.g., infection and CLTI) were included only when there had been a preceding unsuccessful attempt at limb salvage (by open or endovascular means). This study will consider all eligible MLA as discrete episodes.

#### 5.1.2 Exclusion criteria

- Amputation for indications other than CLTI (for example trauma, sepsis, extensive venous ulceration, acute limb ischaemia or chronic pain).
- Patients with insufficient data to calculate a CFS score.
- Minor lower limb amputations (such as digital or transmetatarsal) were excluded. Further, due to anticipated low incidence, hindquarter amputations were also excluded.
- Patients with a CFS score of 9 – these patients are defined as terminally ill irrespective of frailty and therefore we cannot assess its effect on outcome.

#### 5.1.3 Outcomes

- The primary outcome will be all-cause mortality following MLA This will be recorded from the electronic health records. This will be defined as the time from amputation to mortality or censored at the last contact time known alive. The secondary outcomes will include:
- Time to discharge (herein known as length of stay). This will be the time from amputation to discharge alive. Any patients that die in hospital will be censored at the date of death.

#### 5.1.4 Key exposure

The key exposure under investigation is frailty. Frailty is measured using the Rockwood Clinical Frailty Scale (CFS). This is a 9-point scale that ranges from 1 (very fit) to 9 (terminally ill) evaluating variables like comorbidity, physical function and cognition to generate a standardised score. A score of 9 describes a terminally ill patient who is not otherwise severely frail, for this reason, this study will only consider patients with CFS scores between 1 – 8.

We will recode into these categories to show a description analysis of the distribution of frailty only. This will be coded into three categories: not frail (CFS 1-3), mildly frail (CFS 4-5), and moderately to severely frail (CFS 6-8) for the primary analysis”

#### 5.1.5 Variables

To understand the sample characteristics the following socioeconomic characteristics: Sex at birth; age at amputation, Charlson Comorbidities Index (CCI)^(4)^ and Index of Multiple Deprivation decile.

The following characteristics of the amputation were recorded: type of procedure (elective or emergency), procedure (transtibial, transfemoral and through the knee). Other forms of lower limb amputation will be excluded.

### 5.2 Sampling

The NBT sample will consist of all patients undergoing MLA of the lower limb since November 2017. This date was chosen due to a trustwide change in how notes are filed. This change (sorting by admission rather than volume) makes it possible to find data and via a standardised data collection method.

The NHSGCC sample will consist of all patients undergoing MLA of lower limbs from 1^st^ January 2016 to 10^th^ October 2021.

#### 5.2.1 Size of sample

The only directly comparable study design that we have found was in an e-poster presentation in the British Journal of Surgery also looking at clinical frailty scoring with amputation. This broke down clinical frailty into three groups (non to mildly frail, moderately frail and severely frail). The mortality rate between the non/mildly frail group and the moderately frail group was 27% vs 44%.

The 1-year mortality rate for patients undergoing major lower limb amputation for peripheral arterial disease is estimated at approximately 35%-50%^(2)^. Assuming a 30% mortality in robust and 40% in frail patients (HR=0.7) in a 1:2 ratio (robust:frail), to detect this difference would require 1,000 patients (for a minimum of 335 events of mortality) using a 0.05 significance level and with 90% power.

#### 5.2.2 Sampling technique

We will use consecutive sampling of all patients who have undergone a lower limb amputation for peripheral arterial disease at NBT and NHSGGC over the study period.

### 5.3 Recruitment

The vascular departments at NBT and NHSGCC keep records of all patients who have undergone lower limb MLA with coding for type of operation and indication.

Electronic health care records will be screened to confirm the operation and indication to assess eligibility for inclusion.

#### 5.3.1 Sample identification

Patients will be identified from Vascular department records of amputations over the study period.

#### 5.3.2 Consent

As this is a prospective quality improvement project, we will not have to seek formal consent for inclusion. All data will be pseudonymised. Caldicott Guardian approval was sought prior to receiving access to the NHS GGC dataset.

## 5 STATISTICAL CONSIDERATIONS

### 6.1 Sample characteristics

The sample characteristics will be tabulated partitioned into those that died and those that survive. No hypothesis testing of the baseline amputation sample characteristics will be carried out.

### 6.2 Analysis

#### 6.2.1 Primary analysis

The primary analysis will be carried out using a multivariable Cox proportional baseline hazards regression. The analysis will assess the association between pre-amputation frailty (CFS 1-3, 4-5, 6-8) and mortality. This will be adjusted for the following known clinically important confounders: age (Under 65, 65-79, 80 and older), sex registered at hospital admission (Female, Male), hospital site, type of procedure (Elective, Emergency), Procedure type (transtibial, transfemoral, through knee, other), index of multiple deprivation (IMD 1-3, 4-7, 8-10), Charlson Comorbidity Index (1-2, 3-4, 5+). Whilst the crude hazard ratio (HR) and adjusted hazard ratio (aHR) will be fitted alongside the 95% confidence intervals (95%CI) and p-values, the primary analysis will be interpreted using the adjusted analysis. The baseline proportionality assumption will be assessed visually using Schoenfeld residuals with a log-log plot. Stata V17 (or later) will be used for the statistical analysis.

Patients assessed that are terminally unwell (CFS=9) will be presented alone.

#### 6.2.2 Secondary Analysis

The secondary analysis will be carried out using a multivariable Cox proportional baseline hazards regression. The analysis will assess the association between pre-amputation frailty (CFS 1-3, 4-5, 6-8) and length of stay.

### 6.3 Population under investigation and missing data

Missing data will be explored for pattern missingness.

### 6.4 Subgroup analyses

The following subgroup analyses will the carried out and the comparison between the outcomes for those not frail (CFS 1-3, versus the frail CFS 4-8): Sex; age group; procedure type, deprivation, and CCI.

### 6.5 Interim analysis

An informal analysis will be carried out using only the data from North Bristol NHS Trust.

The analysis will be carried out using a multivariable Cox proportional baseline hazards regression. The analysis will assess the association between pre-amputation frailty (CFS 1-3, 4-5, 6-8) and mortality. This will be adjusted by age, sex, type of procedure, and type of amputation. A statistical significance level of 5% will be used.

## 7 ETHICAL AND REGULATORY CONSIDERATIONS

This prospective study using retrospectively collected data analyses outcomes in patients who have undergone MLA for CLTI. There are no specific risks to participants identified. All data is gained from departmental, hospital records and GP records and will be pseudonymised for analysis. This quality improvement project has been registered with the quality improvement department at NBT and NHSGGC.

### 7.1 Assessment and management of risk

The assessment of risk from this study is low. All information will be taken retrospectively from hospital or GP records.

All records have been entered by healthcare professional and any safeguarding risks should have been raised at the time of recording.

If any concerns are raised from retrospective analysis of notes, they will be escalated to the study head for assessment and actioning.

### 7.2 Amendments

There will be no amendments to this protocol.

### 7.3 Patient & Public Involvement

No patients have been involved in the design of the study.

### 7.4 Data protection and patient confidentiality

Data will be collected by staff currently working at NBT or NHSGGC and stored on secure trust systems. The final database will be collated by the study co-ordinator when pseudonymised. Any data sent for analysis will be anonymised and coded prior to sending.

### 7.5 Access to the final study dataset

Pseudonymised data will only be available to the chief investigator (Philip Braude) and the management group named on the protocol.

In line with many peer-reviewed journal’s policy for data sharing, data sharing may be offered to third parties only on request to the study CI with review by the study sponsor to ensure legitimate academic interest. Data will be shared with anonymous records if deemed appropriate, arranged via data sharing agreement, and transferred using secure systems. If requests originate from outside the EU this will be discussed with the study sponsor.

## 8 DISSEMINIATION POLICY

### 8.1 Dissemination policy

The data will be owned by North Bristol Trust as the sponsor. On completion of the study the data will be analysed, and a study report completed.

It will be disseminated locally via the trust operational update and events, as well as more widely through national and international conference presentations with a vascular surgical, geriatric medicine and frailty theme. A summary of the work will be submitted for peer-reviewed publication.

### 8.2 Authorship eligibility guidelines and any intended use of professional writers

Authorship will take into account all persons involved in the study conception, design, analysis, interpretation and write up. This will be accurately reflected when any papers are submitted for peer-reviewed publication.

## Data Availability

Pseudonymised data will only be available to the chief investigator (Philip Braude) and the management group named on the protocol.
In line with many peer-reviewed journal policies for data sharing, data sharing may be offered to third parties only on request to the study CI with review by the study sponsor to ensure legitimate academic interest. Data will be shared with anonymous records if deemed appropriate, arranged via data sharing agreement, and transferred using secure systems. If requests originate from outside the EU this will be discussed with the study sponsor.

